# Correlation of Suspected COVID-19 Symptoms with COVID-19 Positivity in Children

**DOI:** 10.1101/2022.05.03.22274641

**Authors:** Sanika A. Satoskar, Daniel Hindman, Amyna Husain, Laura Prichett, Oluwakemi B. Badaki, Ann Kane

## Abstract

**Background:** Early in the pandemic, COVID-19 was found to infect adults at higher rates than children, leaving limited data on disease presentation in children. Further understanding of the epidemiology of COVID-19 symptoms among children is needed. Our aim was to explore how symptoms vary between children testing positive for COVID-19 infection versus children testing negative.

**Methods:** Data analysis of symptom prevalence among pediatric patients presenting to emergency departments (ED) in the Johns Hopkins Health System (JHHS) with concern for COVID-19 who subsequently received COVID-19 testing. Inclusion criteria included patients 0-17 years-of-age, ED evaluation between March 15th, 2020 - May 11th, 2020, and those who were ordered for COVID-19 testing. Chart review was performed to document symptoms using ED provider notes. Comparisons were made using chi-squared t-tests and Student’s t-tests.

**Results:** Fever (62.6%) and cough (47.9%) were the most prevalent symptoms among children with suspected COVID-19 infection. Compared to children with a negative COVID-19 test, children who tested positive had higher prevalence of myalgia (21.7% vs 6.0%) and loss of taste/smell (15.2% vs 0.9%). Over half of the children who tested positive for COVID-19 had public insurance (52.2%) and 58.7% of the positive tests occurred among children with Hispanic ethnicity.

**Conclusions:** Myalgia and loss of taste/smell were found to be significantly more prevalent among COVID-19 positive children compared to children testing negative. Additionally, children with public insurance and those with Hispanic ethnicity were more likely to test positive, emphasizing the importance of social factors in the screening and decision-making process.

## 1 Introduction

COVID-19 has had widespread effects across the United States (US), including over 70,000,000 cases diagnosed and over 850,000 deaths since the start of the pandemic, as of January 26, 2022 [1]. Early in the pandemic, SARS-CoV-2 was found to infect adults at a much higher rate with more severe manifestations compared to children (0-17 years old) [2-4]. As a result, initial research efforts to understand COVID-19 were focused largely on the adult population, leaving very limited data on disease epidemiology among children.

Recent data shows that COVID-19 infection may present differently in children compared to adults, as seen by differences in the timing of symptom presentation and disease severity [3-6]. It is, therefore, plausible that the prevalence of certain symptoms will vary in the pediatric population compared to adults. Further understanding of the epidemiology of symptoms among children is necessary for guiding decisions regarding testing.

Although many children have asymptomatic infections [6], studies show that children with symptomatic SARS-CoV-2 infections develop symptoms that range in severity across multiple organ systems. These symptoms have been shown to primarily affect the respiratory system, and include cough, sore throat, and shortness of breath [7]. Gastrointestinal (GI) symptoms are also common and include diarrhea and vomiting. Recent evidence shows that the Multisystem Inflammatory Syndrome (MIS-C), a serious condition resulting in systemic inflammation that adversely affects the heart [8], lungs, kidneys and GI organs, is also associated with COVID-19 in children [9-11].

However, an analysis to further explore the association between individual symptoms with COVID-19 test positivity has not been performed in children. Assessing whether certain symptoms are more specific for a positive COVID-19 test has utility to the clinical provider, providing a gauge that helps to discern whether any particular symptom is likely to place a patient in one of two groups: COVID-19 illness or not COVID-19 illness. This is crucial as it can help aid in risk stratification and efficient utilization of testing resources in children, which is critical to pandemic control [12-13] especially when resources are scarce, as was evident during the early months of the pandemic.

Given the varying presentations of pediatric COVID-19 illness, we conducted an analysis of symptom prevalence among pediatric patients presenting to emergency departments with concern for COVID-19 who subsequently received COVID-19 testing. We collected data from three pediatric emergency department (ED) facilities in a single health system during the initial stages of the pandemic. Our primary specific aim was to explore how symptoms vary between children testing positive for COVID-19 infection versus children testing negative, which was used to associate individual symptoms to COVID-19 test positivity.

## 2 Methods

### 2.1 Design and Sample

We performed a retrospective secondary data analysis of children from 0 through 17 years of age who presented with possible COVID-19 disease to three Pediatric ED facilities in a single health system – two suburban community-based hospitals (PED 2, PED 3) and an urban designated level 1 pediatric trauma center (PED 1). Relevant encounters took place between March 15^th^, 2020 and May 11^th^, 2020. We included any patient who was ordered for COVID-19 testing during the time period. COVID-19 testing was ordered based on if the patient met the testing algorithm criteria set at the time, and this algorithm changed over the study period as the pandemic evolved. SARS-CoV-2 testing was performed using a PCR assay. Molecular assays used for diagnosis at the time included Xpert Xpress SARS-CoV-2/Flu/RSV (Cepheid), NeuMoDx SARS-CoV-2 (Qiagen), Cobas SARS-CoV-2 (Roche), ePlex Respiratory Pathogen Panel 2 (Roche), Aptima SARS-CoV-2 (Hologic), Accula SARS-CoV-2 assays (ThermoFisher Scientific), and RealStar® SARS-CoV-2 RT-PCR (Altona Diagnostics) [14-17]. Choice of test method was based on what was optimal for fastest result turnaround time given the staffing during the shift and test volumes. ED visits that were transfers from one of the other two facilities were excluded. This study was approved by the Johns Hopkins University Institutional Review Board (IRB00246826).

### 2.2 Data Source

We utilized data collected via a Precision Medicine Analytics Data Commons Platform (PMAP) based on electronic medical records (EMR) [18]. Relevant variables included the date of the visit, the associated pediatric emergency department, vital signs, and patient demographics, including age, sex, race, ethnicity, and insurance. We also included receipt of SARS-CoV-2 testing, results of SARS-CoV-2 testing, isolation orders, acuity measured with the emergency severity index (ESI), presence of influenza-like illness (ILI) symptoms, and the patient’s area deprivation index (ADI) based on the census block.

For every encounter where SARS-CoV-2 testing was obtained, we performed chart review based on a pre-determined protocol to document the presence or absence of symptoms using provider notes in the ED. Pertinent symptoms included fever, headache, sore throat, cough, shortness of breath, myalgias, diarrhea, and loss of taste or smell. We used REDCap for data entry. All charts that were flagged for questions were reviewed by a single senior investigator to resolve any ambiguity. Missing data were not imputed, however there were very little missing data. Out of the total 516 patients, 18 patients had missing data for some of the variables and these patients were excluded from the analysis involving those variables.

### 2.3 Data Analysis

We analyzed data using Microsoft Excel for Mac 2021 and StataIC 16.1 (StataCorp. 2019. *Stata Statistical Software: Release 16*. College Station, TX: StataCorp LLC). The prevalence of each pertinent symptom was found by dividing the number of children presenting to the ED with that symptom over the total ED visits in our cohort during that time period. The prevalence rate of each symptom was then compared between children who tested positive for SARS-CoV-2 to those who tested negative for SARS-CoV-2. Subjective parameters including myalgias, headaches, and loss of taste or smell were analyzed by a subgroup analysis which included only children above 5 years-of-age. We made comparisons using chi-squared t-tests for categorical variables and Student’s t tests for continuous variables.

## 3 Results

We identified 516 patient encounters where providers were concerned for possible COVID-19 and subsequently tested for SARS-CoV-2. There were 46 (8.9%) children who tested positive for SARS-CoV-2 and 470 (91.1%) who tested negative. **Table 1** presents summary descriptive characteristics of the study population.

**Table 1.** Patient Characteristics of those tested for COVID-19. *Note*. NH = non-Hispanic, ADI = area deprivation index. The ^a^p-value is a comparison of patient characteristics between children who tested positive versus children who tested negative for COVID-19. For continuous variables, means are compared with *t* tests. For categorical variables, proportions are compared with chi-square tests

| Characteristics | Tested (n = 516) | Positive (n = 46) | Negative (n = 470) | <sup>a</sup>p-value |
| --- | --- | --- | --- | --- |
| **Total** | 516 (100.0%) | 46 (8.9%) | 470 (91.1%) | <0.001 |
| **Age (SD)** | 6.64 (5.84) | 8.76 (6.38) | 6.43 (5.75) | 0.01 |
| **Age category** |  |  |  |  |
| **0-13 months** | 137 (26.6%) | 10 (21.7%) | 127 (27.0%) | 0.008 |
| **13 mos-4 years** | 129 (25.0%) | 5 (10.9%) | 124 (26.4%) |  |
| **5-11 years** | 107 (20.7%) | 9 (19.6%) | 98 (20.9%) |  |
| **12-17 years** | 143 (27.7%) | 22 (47.8%) | 121 (25.7%) |  |
| **Female sex** | 262 (50.8%) | 25 (54.4%) | 237 (50.4%) | 0.61 |
| **Race/Ethnicity** |  |  |  |  |
| **NH White** | 149 (28.9%) | 6 (13.0%) | 143 (30.4%) | <.0001 |
| **NH Black** | 171 (33.1%) | 6 (13.0%) | 165 (35.1%) |  |
| **Hispanic** | 111 (21.5%) | 27 (58.7%) | 84 (17.9%) |  |
| **Asian** | 45 (8.7%) | 3 (6.5%) | 42 (8.9%) |  |
| **Other/Unknown** | 40 (7.8%) | 4 (8.7%) | 36 (7.7%) |  |
| **Insurance** |  |  |  |  |
| **Public** | 261 (50.6%) | 24 (52.2%) | 237 (50.4%) | <0.001 |
| **Private** | 208 (40.3%) | 7 (15.2%) | 201 (42.8%) |  |
| **Other** | 24 (4.7%) | 8 (17.3%) | 16 (3.4%) |  |
| **Uninsured** | 23 (4.5%) | 7 (15.2%) | 16 (3.4%) |  |
| **Mean ADI** | 5.13 (2.82) | 5.65 (2.86) | 5.08 (2.82) | 0.19 |

The majority of COVID-19 cases occurred in the 12-17 years age group (47.8%). The mean age among those who tested positive was 8.76 years compared to 6.64 years among those who were negative (p = 0.01). Although the majority of our study population consisted of non-Hispanics (78.5%), 58.7% of positive tests occurred among children with Hispanic ethnicity. The majority of children who tested negative for SARS-CoV-2 were either non-Hispanic Black (35.1%) or non-Hispanic White (30.4%). Over half of the children who tested positive for SARS-CoV-2 had public health insurance (52.2%).

Fever (62.6%) and cough (47.9%) were the most prevalent symptoms among children suspected of SARS-CoV-2. When compared to children with a negative SARS-CoV-2 test, children who tested positive had a significantly higher prevalence of myalgia (21.7% vs 6.0%, p-value = 0.001) and loss of taste or smell (15.2% vs 0.9%, p-value < 0.001), as summarized in **Table 2**. There were no significant differences in the prevalence of sore throat, cough, shortness of breath, and diarrhea symptoms between children testing positive versus negative for SARS-CoV-2.

**Table 2.** Symptom Complaints of those tested for COVID-19. The ^a^p-value is a comparison of symptoms between children who tested positive versus children who tested negative for COVID-19. For categorical variables, proportions are compared with chi-square tests

| Symptoms | Positive COVID test<br>(n= 46) | Negative COVID test<br>(n= 470) | <sup>a</sup>p-value |
| --- | --- | --- | --- |
| History of Fever | 36 (78.3%) | 287 (61.1%) | 0.05 |
| Headache | 11 (23.9%) | 56 (11.9%) | 0.06 |
| Sore throat | 8 (17.4%) | 50 (10.6%) | 0.30 |
| Cough | 24 (52.2%) | 223 (47.4%) | 0.44 |
| Shortness of Breath | 9 (19.6%) | 76 (16.2%) | 0.70 |
| Myalgia | 10 (21.7%) | 28 (6.0%) | 0.001 |
| Diarrhea | 2 (4.3%) | 56 (11.9%) | 0.25 |
| Loss of taste or smell | 7 (15.2%) | 4 (0.9%) | <0.001 |

## 4 Discussion

In this retrospective secondary data analysis utilizing EMR data and chart review, we found that among children who present with suspected COVID-19 infection, fever and cough are the most prevalent symptoms. However, we find that symptoms of myalgia and loss of taste or smell are associated with COVID-19 positivity in our pediatric patient population.

Although many studies have documented a variety of symptoms, including these [6-7], fever and cough have been reported as two predominant symptoms associated with SARS-CoV-2 infection in children [7-8]. Fever and cough were also found to be present in the majority of children who underwent testing for COVID-19 infection in our study. However, interestingly, it was myalgia and loss of taste or smell that were significantly associated with a positive COVID-19 test result. Myalgia symptoms have been implicated in many inflammatory conditions [9]. It may be possible that in children suspected of COVID-19 infection, myalgia may be a direct manifestation of the systemic inflammatory state implicated in COVID-19 pathogenesis, and therefore a more specific indicator for positive COVID-19 infection compared to other viral respiratory symptoms alone.

Our study took place during the early months of the pandemic, when testing resources were scarce and limited, and as a result resource allocation and utilization had to be done prudently. Currently, COVID-19 infection rates are increasing in the pediatric population [19-20], and several health systems are starting to face similar testing resource shortages as was evident early in the pandemic [21]. Our study identifies symptoms that are most specific for SARS-CoV-2 positivity in our pediatric study population, which is essential to aid in strengthening COVID-19 test specificity to aid in efficient resource utilization.

In our study population, racial and socioeconomic factors had a strong association with SARS-CoV-2 positivity. Children who had public insurance or no insurance were significantly more likely to test positive for SARS-CoV-2 compared to children with private insurance. Additionally, we found ethnicity to be significantly associated with SARS-CoV-2 positivity where Hispanic children had a higher likelihood of testing positive compared to non-Hispanic children. Although this phenomenon has been documented in adults, our study provides further insight among the pediatric population. This is especially crucial since the urban hospitals under the Johns Hopkins Health System (JHHS) predominantly serve Black and Hispanic populations, mainly from within and surrounding the Baltimore area. Interestingly in our study, most of our study population consisted of non-Hispanic children, however over half of the positive tests occurred in Hispanic children. It has been extensively documented that many children belonging to minority racial and ethnic groups reside in poorer neighborhoods with significantly lower resources [22-24]. These children often live in high-density overcrowded housing, are more reliant on public transportation, and lack preventive health services, all of which may impact their ability to adhere to social distancing guidelines and subsequently increase their susceptibility to COVID-19 exposure [25-26]. Although the increased COVID-19 positivity rates that were observed in Hispanics and in patients with public or no insurance in our study could be in part due to the sociodemographic structure of the surrounding areas, several studies have shown that minority racial groups and those belonging to lower socioeconomic statures were at increased risk for serious and fatal COVID-19 infections throughout the pandemic [27]. Our findings corroborate with this and highlight the importance of incorporating racial and social factors in screening for COVID-19 infection.

Our study has several limitations. First, our data contained sampling and ascertainment bias since SARS-CoV-2 testing was based on symptom presentation. We did not test children at random, rather we tested children based on the presence of certain symptoms, which were pre-determined by our testing algorithms at that time. Initially, these symptoms included only fever and respiratory symptoms. Diarrhea and loss of taste/smell were added late in the study period, which likely explains the observed lower prevalence of those symptoms in our study population. Second, we assessed patient encounters that occurred during the early months of the pandemic. Data on COVID-19 symptoms were just beginning to emerge at that time, which may have played a role in symptom documentation and triage. Third, our study was limited to hospitals that are situated in urban settings in a single state, which limits generalizability. Finally, we were not able to assess social and environmental factors such as household structure, parental occupation, and environmental exposures, all of which may influence disease severity and subsequent symptom presentation.

As evidence continues to grow, our study provides further insight into COVID-19 symptoms among the pediatric population. However, the presence or absence of any single symptom, other than possibly the sudden loss of taste or smell, is not sufficient to guide clinical decision-making in testing for SARS-CoV-2 among pediatric patients presenting to emergency facilities. When considering testing for COVID-19, clinicians should be attuned to other factors that may predispose patients to increased risk, such as social determinants of health and patients’ social habits. Additionally, given that COVID-19 infectivity is growing in the pediatric population with the addition of newer variants as compared to the start of the pandemic, it is essential that clinicians maintain a low threshold for testing children with a variety of symptoms, allowing for improved disease detection and aiding the prevention of subsequent spread.

## Data Availability

Data cannot be shared publicly because it contains patient data. Data are available from the Johns Hopkins Institutional Data Access/Ethics Committee (contact via 410-955-3008) for researchers who meet the criteria for access to confidential data.

## 6 Conflict of Interest

The authors declare that the research was conducted in the absence of any commercial or financial relationships that could be construed as a potential conflict of interest.

## 7 Author Contributions

SS conceptualized and designed the study, analyzed, and interpreted the data, drafted the manuscript, and critically revised the manuscript for intellectual content.

DH, AH, LP, OB, and AK conceptualized and designed the study, acquired, analyzed, and interpreted the data, critically revised the manuscript for intellectual content, provided administrative, technical, and material support for the study, and provided overall study supervision.

All authors read and approved the final manuscript.

## 8 Funding

This project was funded by the Emergency Medicine Foundation COVID-19 Research Grant

## 10 Abbreviations

US: United States
ED: emergency department
COVID-19: coronavirus disease 2019
SARS-CoV-2: severe acute respiratory syndrome coronavirus 2
MIS-C: multisystem inflammatory syndrome in children
ILI: influenza-like illness
PMAP: Precision Medicine Analytics Data Commons Platform
ESI: emergency severity index
ADI: area deprivation index EMR = electronic medical record
GI: gastrointestinal
ILI: influenza-like illness

## 11 Data Availability Statement

The dataset generated and analyzed for this study can be obtained from the corresponding authors upon request.

## Notes

### Competing Interest Statement

The authors have declared no competing interest.

### Funding Statement

DH and AH received an Emergency Medicine Foundation for COVID-19 grant. The funders had no role in study design, data collection and analysis, decision to publish, or preparation of the manuscript.

### Author Declarations

The Johns Hopkins Institutional Review Board (IRB00246826) approved this study.

